# Testing the effects of nudging for reduced salt intake among online food delivery customers: a research protocol for a randomized controlled trial

**DOI:** 10.1101/2021.06.30.21259691

**Authors:** Beisi Li, Ying Cui, Chao Song, Wenyue Li, Jun Nakagawa, Paige Snider, Ailing Liu, Ying Long, Gauden Galea

## Abstract

**Background:** Chinese people on average consume almost twice as much salt as recommended by the World Health Organization. In recent years, dining out and ordering food online are increasingly popular, especially for urban residents. The aim of this study is to evaluate the effectiveness of different settings on a digital food delivery App in nudging consumers towards reduced salt options through a randomized controlled trial in China.

**Methods and Analysis:** This is a randomized controlled trial with matched restaurants randomized to five parallel intervention groups plus a control group. Participating restaurants are recruited via open invitation and targeted invitation on a voluntary basis and are free to withdraw from the study at anytime. Each enrolled restaurant can select 1-3 of their most popular dishes to participate in this study. The recruitment ends at the end of June 2021. As of June 30, 285 restaurants enrolled for intervention groups and successfully completed interface set-up requirements. The primary outcome of this study is to investigate the differences in customer ordering behaviors regarding salt preference that result from changing the default settings and/or in combination with health messages before placing the order. The primary outcome will be measured by the difference between the number of regular salt orders and the number of reduced salt orders amongst the five intervention groups, and between each intervention group and the control group. We will collect order data at the end of the 2-month study period from the food delivery App. The secondary outcome is to measure if reduced the salt version of the participating dishes has less salt content than the regular version. The secondary outcome will be measured by lab testing salt content of randomly sampled dishes during the study period. In addition, we will also conduct pre- and post- intervention surveys with participating restaurants to assess their knowledge, attitude, and practice regarding salt reduction, and their perceptions on how such intervention affects their business, if at all. We will not include findings from the pre- and post-intervention interviews as an outcome but will use them to inform future restaurant- based salt reduction promotions.

**Discussion:** The study will test whether changing in the choice architecture on the digital food ordering platform will promote healthier ordering behavior among consumers. Results on whether user interface modifications can promote purchases of reduced salt dishes may provide evidence to inform future sodium reduction strategies and health promotion interventions on online food ordering platforms, with the potential to apply to offline dining settings. The results may also inform current government efforts to roll out national guidelines on promoting nutrition labeling by restaurants. Despite these strengths in study design, securing the agreement of the food delivery App, recruiting individual restaurants and maintaining compliance to the interface set up through the period of the study proved to be and remains challenging.

**Trial Registration:** Registered at the Chinese Clinical Trial Registry at http://www.chictr.org.cn/searchprojen.aspx (No. ChiCTR2100047729)

**Protocol version:** Version MAY112021.01

**Strengths and Limitations:**

- This study presents an innovative and timely intervention package for promoting salt reduction for online meal ordering platform.
- Our study is one of first in China and globally that tests nudging interventions on online food ordering behavior on a large-scale commercial platform and in a real-world setting.
- Recruiting restaurants and ensuring their compliance remain challenging.

## INTRODUCTION

### Background

Most people in China consume too much salt on average. The most recent national survey conducted between 2015 and 2019 found that the mean daily salt intake from cooking sources was 9.3g per person, not including the amount of salt that was consumed from other sources, i.e. pre-packaged foods and restaurant dishes[1]. This is almost double the recommended daily intake of 5g of salt per day, according to both WHO and the Healthy China Action Plan 2019-2030[2-3]. Too much salt in the diet means too much sodium, a major risk factor for developing high blood pressure and cardiovascular diseases, mainly stroke and coronary heart disease. There are 270 million adults in China with high blood pressure[3]. Cardiovascular disease is the number one killer in China, causing two in five deaths in China[4]. Trends indicate that as China urbanizes and incomes rise, food consumption away from home has been increasing as a source of food for city dwellers[5-6]. Online meal delivery services are becoming a popular source of prepared meals in China[7]. Online platforms typically provide little to no dietary information to guide consumer decisions.

This study investigates whether low-costs ‘nudges’ on an online food ordering platform can encourage consumers to choose low sodium options. Nudges are defined as “any aspect of the choice architecture that alters people’s behavior in a predictable way (1) without forbidding any options or (2) significantly changing their economic incentives”[8]. Unlike traditional educational messages that require consumers to deliberately process information, nudging relies on subtle changes at the point of decision-making to make it more effortless and convenient to engage in the healthier choice.

The primary focus of this study is to measure the effectiveness of nudge interventions on Eleme, a popular food delivery App in China, to encourage the consumer to choose reduced salt options at the point of purchase. Findings from this study will inform the growing number of field experiments demonstrating how nudges can promote healthy eating. The secondary focus is to evaluate whether the salt content in the reduced salt version of dishes is less than in dishes with regular salt content.

#### Review of existing studies

Traditionally, providing health information such as signage at point of sale and educational or health literacy campaigns are the most common approaches for promoting healthy eating. However, review of existing evidence showed mixed findings regarding information’s role in promoting heathy eating behaviors. In a meta-analysis, Cadaro and Chandon[9] found that affective and behavioral nudges did significantly better than informational nudges. In another systematic review, Harbers et al.[10] examined 75 studies that showed mixed results regarding information nudges. They found in general information nudges had moderately positive effects, but the effects can be heterogeneous across studies.

A few studies have applied nudges to affect consumers food choices. In these studies, researchers deployed intervention strategies, such as position nudge in menu design to affect consumers’ food choices and measured the outcomes by caloric intake[11-14], fat[11,13], sugar[11], sodium[11,12] or salt, fruit and vegetables[15], and vegetarian[16,17]. Most of the studies were conducted on online food ordering platforms for workplace and school canteens[11,15], rather than on commercial food ordering systems that serve the whole population. Hence, the findings are not representative for the population. Some studies were conducted in a controlled lab environment rather than in the natural setting[12,14,18], which renders the findings less powerful as recommendations for real world practice. While altering the default has been used to encourage healthy eating for school lunch programs, nutrition assistance programs, and hospital cafeterias[19,20], few studies have examined healthy defaults as applied to food selection on digital ordering platforms. Other studies have shown that front-of-package sodium labelling using traffic light and warning labels significantly reduced consumption of sodium[21,22].

One of the most powerful and simplest nudges is setting the default. Behavioral science research has consistently demonstrated that people tend to accept the default choice provided to them because of the additional cognitive investment required to opt-out of the status quo[23,24]. Changing the default from opt-in to opt-out option has shown to be highly effective in increasing participation in flu vaccines, organ donation, and retirement saving programs[25,26].

To the best of the authors’ knowledge, no study to date has investigated the effect of inexpensive nudges to lower consumption of sodium on an online meal delivery platform in the real-world setting. Moreover, studies that applied nudges to promote healthy diet were largely conducted in high-income countries, and none were found from developing countries.

## METHODS AND ANALYSIS

### Trial Design

This is a randomized controlled trial with matched restaurants randomized to 6 parallel groups, including 5 intervention groups and 1 control group, without cross-over. The trial is in factorial design. The intervention includes three treatments, including setting up a sub-menu on the menu interface for ‘Reduced Salt’ and ‘Regular’ options, sub-menu defaults on either ‘Reduced Salt’ or ‘Regular’ options, and a health message to ‘eat no more than 5g salt per day’, the standard health education message used by the government. Each group is assigned to one of the treatments or one combination of selected treatment (See Fig.1 Study Design). Interactions are anticipated between health message and default and between sub-menu, message and default.

**Fig. 1.**
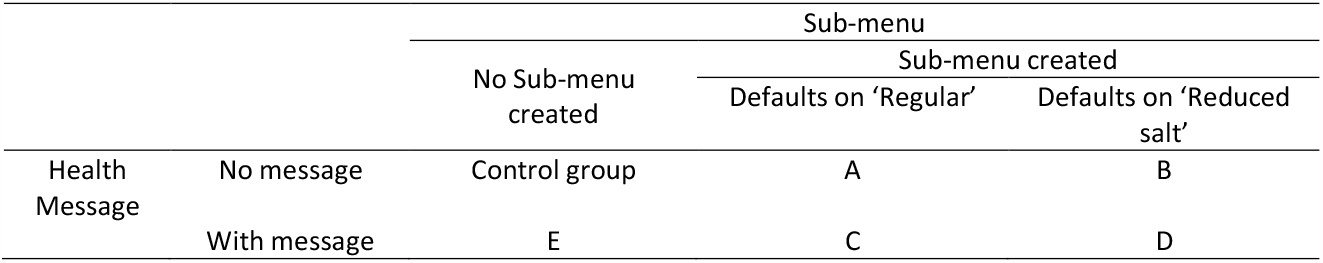
Study Design

### Restaurant Recruitment

Restaurants are recruited on a voluntary basis. We use two different methods to engage restaurants. The restaurants join by either responding to Eleme’s public invitations or by responding to targeted invitations through the research teams’ and Eleme’s existing networks. Since participation is voluntary, restaurants are free to leave the project at any time.

In December 2020, Eleme issued an open invitation in seven cities through their internal communication channel with individual restaurants. In March 2021, China CDC and Eleme started targeted invitations through their existing partnerships in three cities (Chengdu, Taizhou, and Taiyuan) and with two restaurant chains (Yunhaiyao and Ziguangyuan). As of June 30, 285 restaurants in total completed the interventional set-up. Among them, 97 were recruited from Eleme’s open invitation, and 188 were by targeted invitation via the team’s existing networks (See Fig.2 Restaurant Recruitment). Each restaurant is asked to select 1-3 of their most popular dishes to participate in this study.

**Fig. 2.**
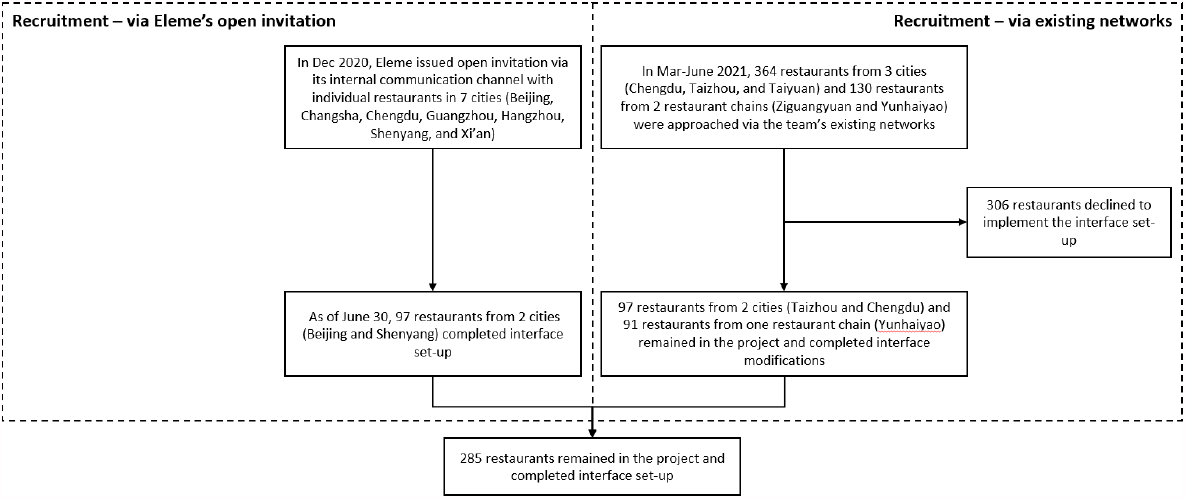
Restaurant Recruitment

We will include data in the study on all restaurants that sign up to participate and complete interface set-up during the intervention period. We do not invite any restaurants that only serve unsuitable dishes for sodium reduction, e.g., pastries, sugary beverages.

We carry out two approaches to improve adherence. One is to target chain restaurants and leverage existing government and business partnerships. We have set up online chat groups with owners or managers of participating restaurants in Taizhou and Chengdu, and with the chain restaurants, respectively, to answer questions, provide guidance, and supervise for compliance. In addition, and at the recommendation of China CDC and Eleme, we also plan to give RMB100 (approximately USD15) when the trial completes to incentivize participation and adherence to the first 200 restaurants recruited by Eleme that successfully complete interface set-up. Eleme also provides a marketing promotion opportunity to promote the restaurants on its homepage after the study is completed as additional incentive to the restaurants. We deploy these two measures to create a sense of obligation to help bind them to the study. The research team will check restaurants’ interface once a week to monitor compliance and to identify any restaurant that fails to follow the protocol.

### Randomization

In order to maximize the comparability among restaurants across groups, we allocate all participating restaurants by randomized block design.

For restaurants recruited by open invitation, we firstly sort all restaurants by food categories and then rank all restaurants within each food category by the number of monthly orders. Based on the ranking, we put restaurants into one block stratified by food category and the order volume. After that, we allocate the restaurants from each block randomly into the five treatment groups. Eleme provided the research team with the order volume data and the research team was responsible for generating the allocation sequence and allocating restaurants into the different groups. To establish comparators, Eleme will extract the ordering data from restaurants that showed intention to participate but do not complete the interface set-up and with similar attributes to those participating.

For those restaurants recruited via targeted invitation via China CDC’s existing networks, researchers initially sorted participating restaurants into blocks stratified by food category, and then allocated the restaurants randomly into the five treatment groups and to the control group.

#### Assignment of Responsibilities

Eleme and the research team were responsible for recruitment. The research team was responsible for allocation of restaurants into intervention groups. Eleme follows up with restaurants after allocation for staff training and monitoring compliance. The statistician who does allocation is not blinded, because the statistician is not involved in direct communication with the restaurants. The participating restaurants are not blinded given the nature of the intervention.

### Intervention

#### Intervention description

In this study, the intervention groups include A) Default on ‘regular salt’ option; B) Default on ‘reduced salt’ option; C) Health reminder and default on ‘regular salt’ option; D) Health reminder and default on ‘reduced salt’ option; and E) Health reminder only (see Fig.1 Study Design). The design of the study will allow researchers to understand the treatment effect of the three treatments of default, sub-menu, and health reminders, and the different combination of the treatments.

The interventions are to test the following hypotheses:

**H1**: Selections which default consumer into the ‘Reduced Salt’ option at sub-menu will result in a higher number of reduced salt orders compared to selections in which consumers are defaulted into the ‘Regular’ option at sub-menu.

**H2**: Combining a health message with a default on ‘Reduced Salt’ will result in a higher number of reduced salt orders compared to ‘Reduced Salt’ default only.

**H3**: When setting up a sub-menu for salt level, regardless of the default setting or the use of a health message, the restaurants will receive a higher number of reduced salt orders compared to restaurants that do not set up a sub-menu at all.

**H4**: A health message alone will not result in higher number of orders with reduced salt requests.

**H5**: When consumers order the reduced salt dishes, including ordering placed via the sub-menu or by leaving a request for less salt in the comment box, the restaurants will add less salt when they prepare the reduced dishes.

#### Ordering page set-up

For Intervention Group A-D, a sub-menu offering ‘Regular’ and ‘Reduced salt’ options will be created under each of the participating dishes, where consumers are prompted to confirm their selection. In Group A and C, the default choice is on ‘Regular’. Customers who prefer less salt will have to actively and consciously change the selection from the default setting of ‘Regular’ to ‘Reduced salt’. In Group B and D, the default setting is on ‘Reduced salt’. For Group E, no such sub-menu is available and, if consumers would like to ask for less salt, they will have to type in the Comment Box before they place their orders. For Intervention Group C, D, and E, a reminder that daily salt intake should be no more than 5 grams (a salt reduction key message published by the China National Health Commission) is set up and pinned to the top of the ordering page. Participating restaurants are required to keep such set-up throughout the study period. For the Control Group, neither the salt-reduction sub-menu nor salt-reduction message reminders are set up.

#### Training for restaurant staff

Nutrition experts at China CDC developed a series of training materials, including a manual and videos, to encourage and guide chefs to reduce the use of salt and salty condiments when preparing dishes. Staff at the participating restaurants are required to watch the training video online before the intervention started. This training mainly focused on the following aspects: ‘project introduction’, ‘why reduce salt’, and ‘practical skills in reducing salt for restaurant cooking.’

#### Pre-and Post-intervention survey with restaurant

We will conduct surveys with selected participating restaurants to understand their knowledge, attitude, and practice regarding salt reduction, and their perceptions on how such intervention affects their business, if at all, before and after the trial period. The surveys will be administered by an online survey platform before and after the intervention as baseline and follow-up, respectively. All participating restaurants are required to finish the online surveys. Questions from the survey and the interview will include (1) basic information about the restaurant; (2) knowledge, attitude, and practice related to reducing salt; (3) perception on how the project will/did affect their business (see Annex 1 for survey questionnaires). Findings will not be included as an outcome assessment but used to inform future restaurant-based salt reduction strategies. The pre-and post-intervention survey will provide us with first-hand insights to understand if such effects exist and to what extent, together with using ordering data we will collect at the end of the study.

Given the ongoing COVID-19 pandemic, we have minimized in-person human contact to carry out the study. Engagement and regular communications with restaurants are conducted via virtual communications tools, such as instant messaging Apps. For restaurant staff training, we disseminated written training materials and pre-recorded videos to participating restaurants to learn independently, instead of organizing in-person training sessions. Surveys with restaurant staff are conducted online via an online survey platform. The research teams have also been following physical distancing recommendations and having regular communications virtually.

### Planned Outcome Assessment

#### Primary outcome

The primary outcome of this study is to measure the differences in consumer behavior regarding salt preference that result from changing the default settings and/or in combination with health messages before placing their food order on the digital platform. We will measure the outcome by comparing the difference between the volume of regular salt ordering and the volume of reduced salt ordering amongst and between the five intervention groups and with the control group.

We will assess the primary outcome at the end of the 2-month intervention period. We will collect the number of regular salt orders and the number of reduced salt orders from each intervention group and the control group. (Note: reduced salt order refers to orders from consumers who chose the reduced salt option in the sub-menu of selected dishes in Intervention Group A-D, and those from consumers who left comments asking for reduced salt in Intervention Group E and the control group). Then we will compare the number of regular salt orders and the number of reduced salt orders amongst different intervention groups, and between each intervention group and the control group. All restaurant ordering data will be acquired from Eleme.

#### Secondary outcome

The secondary outcome is to evaluate whether the salt content in the reduced salt version of dishes across intervention groups is less than in dishes with regular salt content. The secondary outcome will be measured by the difference of salt content between the reduced salt version and the regular version of the participating dishes. To do so, we will randomly order the regular salt versions and reduced salt version of participating dishes during the trial period and test and compare their salt content in a laboratory. A sampling plan of sodium content measurement has been prepared. We will also use the secondary outcome assessment as a mean for validation to monitor if the restaurants indeed put less salt in the reduced salt version. If the test result shows the difference in salt content is less than 10%, we will contact the restaurant to ensure compliance.

### Planned Statistical Analyses

We expect to see different behaviors and choices made by restaurants. For example, some restaurants could choose to set up the user interface for a different intervention group than the one they were assigned. This could potentially limit randomization and introduce bias. To account for such issues, we plan to conduct data analyses on restaurants by their original randomized allocation and by groups they are actually in. The proposed approach will also allow for real-world uncertainties and examine how the results of the study might change under different circumstances, which will increase the level of sensitivity of our trial.

#### Primary outcome analysis

The effect of different treatments will be analysed if needed using chi-square test, analysis of variance (ANOVA), and regression models. We will use different data points and analysis to assess the effectiveness of different treatments and treatment combinations (see Fig.3 Planned Primary Outcome Assessment).

**Fig. 3.**
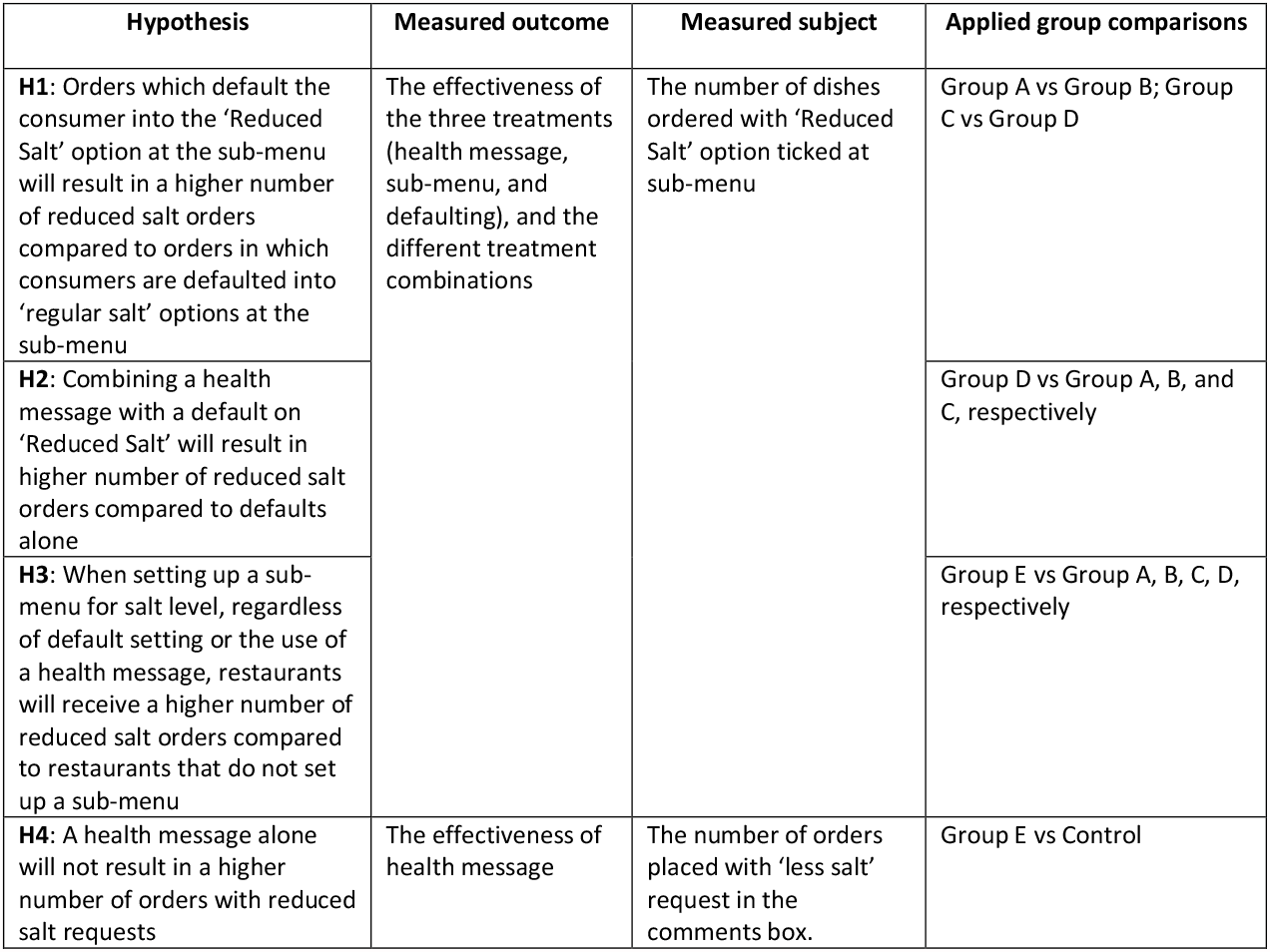
Planned Primary Outcome Assessment

#### Comparability Analysis

To support primary outcome assessment, we plan to conduct a comparability analysis on several variables at baseline and at the end of the trial to account for the imbalance across groups. First, we plan to use difference-in-difference analysis to examine if the trend of changes in number of orders with reduced salt requests are even across groups. Second, we will also choose food type as the variables for comparison, based on the assumption that people’s impression of the saltiness of one type of cuisine could become a reason for reduced salt request. Third, because the consumers’ social economic attribute may influence their salt reduction choice, we also consider comparing restaurant location and price range indicated to consumers on the App, two proxies that suggest consumers’ socioeconomic status. The actual comparability analysis will be conducted based on data access and availability.

#### Secondary outcome analysis

For H5 to monitor if the reduced sodium version of an ordered dish indeed contains less sodium, we will randomly order the participating dishes in regular salt and reduced salt versions and test the salt content in the laboratory during the intervention period.

We assumed the SD of 1 g/100 g of dish for the sodium content and expected to achieve 80% power (with two-sided alpha=0.05) and to detect a change in sodium content by 0.5g/100g in one dish. Meanwhile, the sodium content with regular salt is assumed to be 5g/100g. The correlation coefficients of the sodium content in regular salt and reduced salt group was 0.1. The minimum sample size of each layer was estimated by paired means test. After calculation by SAS V.9.4, the sample size in each layer is about 60 pairs of dishes. The dishes will be randomly selected from the five treatment groups (Group A-E). 60 regular dishes in each layer will be ordered in the early July, and the same version of the reduced salt dishes will be ordered in the late August. The differences in the sodium content in regular salt and reduced salt version will be estimated according to two independent samples or paired samples by Chi-squared and t-tests.

#### Data management and confidentiality

This study does not involve individuals’ personal information. This study is to primarily measure the number of digital orders from participating restaurants in different intervention and control groups. Eleme will share order information after pseudonymization at the end of the study period. No order information will be traced back to individual customers in this study. Eleme’s privacy terms and policies are written such that no additional consent will be required from individual customers when the App is sharing pseudonymized information with academic institutes for public interest purposes or academic research (Eleme Privacy Terms and Policies – 3.9). All users have already agreed to the privacy terms and polices, as it is a pre-requisite for use of the App.

### Project Timeline

Restaurant recruitment and allocation are to be completed at the end of June 2021. Order data from July 1 will be counted as long as the restaurants have applied interface set-up. For secondary outcomes, China CDC plans to start sampling dishes to monitor salt content during the trial period (See Fig.4 Project Timeline).

**Fig. 5.**
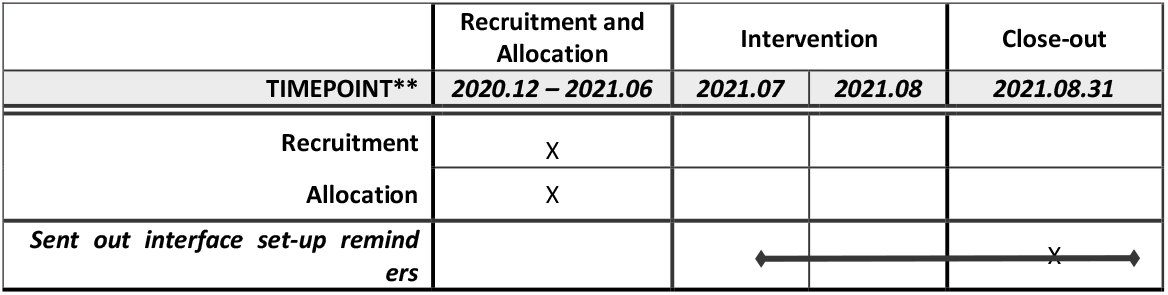
Project Timeline

## DISCUSSION

### Expected outcome and potential impact

China health promotion authorities and research institutes have undertaken efforts for many years to promote salt reduction among customers when dining in restaurants. However, most of the previous approached focused on offline dining environment modifications (e.g., put up health reminders in the restaurants or providing training to the restaurant staff). This study is one of first interventional trials in China and to the authors’ knowledge in Asia that uses behavioral insights and nudge theories to promote customers’ online ordering behaviors towards salt reduction. Existing similar studies are mostly conducted in western countries, e.g., Canada[26], Australia[15], the United Kingdom[13-18], and the United States[14]. Given that western cuisines and taste preferences are different from those in Asia, our study may contribute to the body of references for other Asian countries with similar cooking and eating practices. What’s more, the existing trials that studied online ordering behaviors were either conducted through the form of survey[12,14,27], in school canteens[15,28], or in simulative environments, the limitations of those settings and the relatively smaller sample size may affect effectiveness estimates. In comparison, this trial takes place in the natural setting on a commercial ordering App which covers a considerable collection of commercial restaurants and geographic areas, with a representative sample.

The shift in consumer behavior towards online food ordering has accelerated even more since the COVID-19 pandemic. Additionally, many restaurants require customers to order on their phones or some shared tablets even when they are ordering in a physical establishment. We hope an exploration like ours on how to create and integrate innovative health promotion methods into mobile device-based food ordering platforms may provide evidence and for similar efforts in the future. In China this may contribute to national efforts of reducing salt intake to 5g/day, an objective in the Healthy China 2030 Action Plan. We also plan to disseminate study findings through journal publication.

We have taken the potential impacts on the restaurants and the ordering platform into consideration, aside from study outcome assessments. Both the research team and Eleme agreed that the study will have no appreciable effects on the overall business either for the platform or the restaurants, given the number of orders affected are small compared to the overall volume. Moreover, this sort of experimentation on the ordering interface is regularly conducted by the platform and restaurants themselves as a means of improving sales or improving the efficiency of the conversion process for visitors. This is also in agreement with consumers as they accepted the privacy terms and policies to be able to proceed with placing orders.

## Limitations

Though we tried to minimize characteristic differences during restaurant allocation, the different drop-out rates among groups may cause unevenness across groups. The unevenness in the food category of restaurants across groups may also affect the effectiveness estimates. It is possible that restaurants serving certain type of cuisines may be more likely to see more consumers’ requests for reduced salt because of the nature of the dish rather than the type of interventions they saw on the ordering interface. In addition, the research team have limited access to restaurant information, e.g., staff and clientele profile, etc. The limited access to restaurant information makes it difficult to profile the restaurants and investigate if there are any factors with the restaurants and their clientele would interact with and affect the effectiveness estimates.

## Data Availability

Due to commercial concerns of the food delivery ordering platform, sharing of individual order data used to support the conclusions is not available.

## Copyright

I, the Submitting Author has the right to grant and does grant on behalf of all authors of the Work (as defined in the author license), an exclusive license and/or a non-exclusive license for contributions from authors who are: i) UK Crown employees; ii) where BMJ has agreed a CC-BY license shall apply, and/or iii) in accordance with the terms applicable for US Federal Government officers or employees acting as part of their official duties; on a worldwide, perpetual, irrevocable, royalty-free basis to BMJ Publishing Group Ltd (“BMJ”) its licensees.

The Submitting Author accepts and understands that any supply made under these terms is made by BMJ to the Submitting Author unless you are acting as an employee on behalf of your employer or a postgraduate student of an affiliated institution which is paying any applicable article publishing charge (“APC”) for Open Access articles. Where the Submitting Author wishes to make the Work available on an Open Access basis (and intends to pay the relevant APC), the terms of reuse of such Open Access shall be governed by a Creative Commons license – details of these licenses and which license will apply to this Work are set out in our license referred to above.

## Ethics Approval

All procedures involving human participants were approved by the Ethics Committee of the National Institute for Nutrition and Health of the Chinese Center for Disease Control and Prevention. Restaurant managers/staff who are interviewed for the survey will sign an informed consent form. Approval reference No. 2021-019.

## Consent to Participate

The restaurant owner/staff who participated in the survey and/or the interview have signed an informed consent from the research team. Restaurants gave their consent to participate by joining the online chat groups created for compliance monitoring and following instructions to set up their user interface.

## Declaration

The Corresponding Author has the right to grant on behalf of all authors and does grant on behalf of all authors, an exclusive license (or non exclusive for government employees) on a worldwide basis to the BMJ Publishing Group Ltd to permit this article (if accepted) to be published in BMJ editions and any other BMJPGL products and sublicences such use and exploit all subsidiary rights, as set out in our license.

## Authors’ contribution

PS and CY conceived the project. All authors participated in initial study design and implementation, CY, LBS, JN, SC, and LWY finalized the study design, instrument development, and implementation plan. SC, LWY, CY, and LBS facilitated restaurant engagement. LWY oversees compliance monitoring and SC is responsible for supervising restaurants for interface set-up and validation. LBS wrote the first draft of the manuscript, and GG, CY, JN, PS, SC, and LWY contributed to the draft. LBS, JN, CY, PS, GG, SC, and LWY contributed to the finalization of the study protocol and approval of the final manuscript. Guarantor: Li Beisi

## Transparency declaration

The lead author affirms that the manuscript is an honest, accurate, and transparent account of the study being reported; that no important aspects of the study have been omitted; and that any discrepancies from the study as planned and registered have been explained.

## Patient and public involvement

Patient and/or the public were not involved in the design, or conduct, or reporting, or reporting, or dissemination plans of this study.

## Acknowledgements

The authors would like to thank the following individuals from Eleme for their invaluable services to implement the project on their platform: Shuhan Zhang, Hong Miao, and Wang Shuang.

## ANNEX 1: SURVEY QUESTIONNAIRES WITH RESTAURANTS

*(The actual questionnaire is in Chinese. This English translation is for internal reference only*.)

1. Would you like to participate in this salt reduction promotion project?
  A. Yes
  B. No
2. Does your restaurant have materials available to the customer on the physical premise related to salt reduction (such as salt reduction posters, table posts, etc.)? (if A is chosen, please jump to question 4)
  A. Yes
  B. No
  C. Not sure (if A is chosen, please jump to question 4)
3. Who provide these materials?
  A. Government or community provided, and restaurants are required to post
  B. Government or community provided, and restaurants can voluntarily post
  C. Self-made and posted
4. Have chefs and waiters been trained to reduce salt?
  A. The chefs only
  B. The waiters only
  C. Both the chefs and the waiters
  D. Neither the chefs nor the waiters
  E. Not sure
5. Would your restaurant be willing to develop reduced salt dishes?
  A. Yes
  B. No
6. Would you like to actively offer customers reduced salt dishes or reduced salt options on the menu? (if A is chosen, please jump to question 8)
  A. Yes
  B. No
7. Why you are reluctant to offer reduced salt dishes? (Choose all that apply)
  A. Concerned about changing the taste of dishes
  B. Worried that the chef can’t make reduced salt dishes
  C. Worried that the customers will not be satisfied with the dishes
  D. Worried about extra costs
  E. No request for less salt from the customers
  F. Too much trouble
  G. Other_________
8. Why are you willing to offer reduced salt dishes? (Choose all that apply)
  A. Customers demand less salt
  B. Reduce costs
  C. Improve restaurant image (publicity)
  D. The government request or advocacy
  E. Good for health
  F. Other______
9. In the long run, do you think promoting salt reduction will have a positive impact on the restaurant?
  A. Yes
  B. No
  C. Not sure
10. Will waiters at your restaurant premises voluntarily ask the customers if they would like to have less salt in them?
  A. Yes
  B. No
  C. Not sure
11. Will the salt be reduced when the consumer requires less salt? (if A is chosen, please jump to question 12)
  A. Yes
  B. No
  C. Not sure
12. Why do you think the staff at your restaurant will not reduce salt even if the customers have requested? (Choose all that apply)
  A. Concerned about changing taste of dishes
  B. Worried that the chef does not know how to make the dish with less salt
  C. Worried that the customers are not satisfied with the dishes afterwards
  D. Other___
13. How do the chefs make reduced salt dishes? (Choose all that apply)
  A. Add less salt during preparation
  B. Add fewer condiments containing salt (such as soy sauce)
  C. Use other ingredients such as onion, ginger and garlic instead
  D. Add salt right before serving
  E. Other____
14. Which of the following condiments will be provided either at the table or for delivery? (Multiple)
  A. None
  B. Salt
  C. Vinegar
  D. Soy sauce
  E. Flour sauce /chili sauce / soybean sauce / yellow sauce
  F. Peanut butter / salad sauce
  G. Pickled mustard / pickled leaf mustard
  H. Other____
15. Do you know what elements of the salt have health effects?
  A. Ca
  B. Na
  C. K
  D. I
  E. Not sure
16. Which of the following condiments contain higher salt? (Choose all that apply)
  A. Vinegar
  B. MSG/Chicken powder
  C. Soy sauce
  D. Flour sauce / chili sauce / soybean sauce / yellow sauce
  E. Preserved Tofu/ stinky tofu
  F. Peanut butter / salad sauce
  G. Pickled mustard / pickled leaf mustard
  H. Not sure
17. Which of the following diseases is associated to high salt intake?
  A. No association
  B. Hypertension
  C. Heart disease
  D. Stroke
  E. Kidney disease
  F. Osteoporosis
  G. Asthma
  H. Diabetes
  I. Obesity
  J. Cancer (such as gastric cancer)
  K. Not sure
  The following two questions will be added to the post-intervention survey:
18. Do you think the intervention has affected the number of online ordering at your restaurant, compared to the number of before the intervention?
  A. No impacts
  B. The restaurant received more online orders
  C. The restaurant received fewer online orders
19. Do you see more online orders with low salt request during the intervention, compared to the number of before the intervention?
  A. Yes
  B. No

## REFERENCES

[1] The State Council Information Office of the People’s Republic of China. Report on the Nutrition and Chronic Diseases Status of Residents in China [Internet]. 2020 Dec. Available from: http://www.scio.gov.cn/xwfbh/xwbfbh/wqfbh/42311/44583/wz44585/Document/1695276/1695276.htm.

[2] World Health Organization [Internet] Salt reduction [Internet]. 2020 Apr. Available from: http://www.who.int/news-room/fact-sheets/detail/salt-reduction.

[3] National Health Commission. Healthy China Action Plan 2019-2030 [Internet]. 2019 Jul. Available from: http://www.nhc.gov.cn/guihuaxxs/s3585u/201907/e9275fb95d5b4295be8308415d4cd1b2.shtml.

[4] Ma L, Chen W, Gao R, et al. China cardiovascular diseases report 2018: an updated summary. J Geriatr Cardiol. 2020;17(1):1–8.

[5] Dong X, Hu B. Regional Difference in Food Consumption Away from Home of Urban Residents: A Panel Data Analysis. Agriculture and Agricultural Science Procedia. 2010;1:271–7.

[6] Zhai F, Du S, Wang Z, et al. Dynamics of the Chinese diet and the role of urbanicity, 1991-2011. Obes Rev. 2014;15 Suppl 1:16–26.

[7] China Internet Network Information Center. The 45th China Statistical Report on Internet Development[Internet]. 2020 Apr. Available from: http://www.cac.gov.cn/2020-04/27/c_1589535470378587.htm

[8] Thaler RH, Sunstein CR. Nudge: improving decisions about health, wealth, and happiness. New Haven: Yale University Press; 2008. 293 p.

[9] Cadario R, Chandon P. Which Healthy Eating Nudges Work Best? A Meta-Analysis of Field Experiments [Internet]. Rochester, NY: Social Science Research Network; 2018 Sep [cited 2021 Jun 22]. Report No.: ID 3090829. Available from: https://papers.ssrn.com/abstract=3090829

[10] Harbers MC, Beulens JWJ, Rutters F, et al. The effects of nudges on purchases, food choice, and energy intake or content of purchases in real-life food purchasing environments: a systematic review and evidence synthesis. Nutr J. 2020;19:103.

[11] Delaney T, Wyse R, Yoong SL, et al. Cluster randomised controlled trial of a consumer behaviour intervention to improve healthy food purchases from online canteens: study protocol. BMJ Open. 2017;7:e014569.

[12] Byrd K, Almanza B, Ghiselli RF, et al. Adding sodium information to casual dining restaurant menus: Beneficial or detrimental for consumers? Appetite. 2018 Jun 1;125:474–85.

[13] Theis DRZ, Adams J. Differences in energy and nutritional content of menu items served by popular UK chain restaurants with versus without voluntary menu labelling: A cross-sectional study. PLoS One. 2019;14:e0222773.

[14] Kim M. The effect of 99-ending calories and anticipated guilt on restaurant menu development strategy. International Journal of Hospitality Management. 2020;89:102570.

[15] Wyse R, Gabrielyan G, Wolfenden L, et al. Can changing the position of online menu items increase selection of fruit and vegetable snacks? A cluster randomized trial within an online canteen ordering system in Australian primary schools. American Journal of Clinical Nutrition. 2019;109:1422–30.

[16] Sparkman G, Weitz E, Robinson TN, et al. Developing a Scalable Dynamic Norm Menu-Based Intervention to Reduce Meat Consumption. Sustainability. 2020;12:2453.

[17] Vaan JM, Steen T,Müller BCN. Meat on the menu? How the menu structure can stimulate vegetarian choices in restaurants. J Appl Soc Psychol. 2019;49:755–66.

[18] Bacon L, Krpan D. (Not) Eating for the environment: The impact of restaurant menu design on vegetarian food choice. Appetite. 2018;125:190–200.

[19] Just D, Price J. Default options, incentives and food choices: evidence from elementary-school children. Public Health Nutr. 2013;16:2281–8.

[20] Thorndike AN, Sonnenberg L, Riis J, et al. A 2-phase labeling and choice architecture intervention to improve healthy food and beverage choices. Am J Public Health. 2012;102:527– 33.

[21] Musicus AA, Moran AJ, Lawman HG, Roberto CA. Online Randomized Controlled Trials of Restaurant Sodium Warning Labels. Am J Prev Med. 2019;57:e181–93.

[22] Taillie LS, Reyes M, Colchero MA, Popkin B, Corvalán C. An evaluation of Chile’s Law of Food Labeling and Advertising on sugar-sweetened beverage purchases from 2015 to 2017: A before-and-after study. Basu S, editor. PLoS Med. 2020;17:e1003015.

[23] Thorgeirsson T, Kawachi I. Behavioral economics: merging psychology and economics for lifestyle interventions. Am J Prev Med. 2013;44:185–9.

[24] Halpern SD, Ubel PA, Asch DA. Harnessing the power of default options to improve health care. N Engl J Med. 2007;357:1340–4.

[25] Chapman GB, Li M, Colby H, Yoon H. Opting In vs Opting Out of Influenza Vaccination. JAMA. 2010;304:43

[26] Mendoza JE, Schram GA, Arcand J, et al. Assessment of consumers’ level of engagement in following recommendations for lowering sodium intake. Appetite. 2014 Feb 1;73:51–7.

[27] Prusaczyk E, Earle M, Hodson G. A brief nudge or education intervention delivered online can increase willingness to order a beef-mushroom burger. Food Quality and Preference. 2021;87:104045.

[28] Miller GF, Gupta S, Kropp JD, Grogan KA, Mathews A. The effects of pre-ordering and behavioral nudges on National School Lunch Program participants’ food item selection. Journal of Economic Psychology. 2016;55:4–16.

